# Assessment of the impact of the new blister packaging of Biktarvy^®^ (B/F/TAF) on treatment satisfaction of people living with HIV

**DOI:** 10.1101/2024.10.26.24316182

**Authors:** JP Cruz, O Santos, M Rodrigues, R Morais, T Borralho, F Ferreira, A Ferreira, J Albuquerque, P Nogueira, MP Silva, F Antunes

## Abstract

**Background:** Antiretroviral therapy (ART) is highly effective in people living with HIV (PLHIV), but its success depends on treatment satisfaction and adherence. A determinant of satisfaction regards how the medication is delivered to the patient, namely how it is contained (e.g., bottles, blisters, etc). A new packaging of Biktarvy^®^ has been introduced as a monthly blister, aiming to improve satisfaction, facilitate traceability of daily medication, portability, and discretion (reducing stigma associated with ART), and, ultimately, enhance adherence.

**Goals:** The study’s objective was to assess the impact of changing the packaging of Biktarvy® (B/F/TAF) from a standard pill bottle to a monthly blister with a weekly calendar on therapy satisfaction. Additionally, the association between treatment satisfaction and selected patients’ characteristics (e.g., ART duration) was evaluated. A secondary goal was to characterize the association between the change of packaging on patient’s adherence.

**Methods:** This is an observational longitudinal (retrospective and prospective) study with patients following ART for at least six months (ambulatory clinical management) recruited according to a non-probabilistic sequential sampling. Satisfaction was measured at two different moments: at baseline, HIVTSQs were used to assess satisfaction within the previous six months’ use of medication containers (bottles). Six months later, patients filled in the HIVTSQc to assess their perception of satisfaction change with the new packaging (blister). Adherence was assessed by pharmacy medication dispensing at the hospital.

**Results:** The study enrolled 105 patients in two selected centers (102 patients completed the study). Patients were significantly more satisfied (HIVTSQc scores) with ART when using the new Biktarvy® blister pack package. Importantly, gains of ART satisfaction were higher among those less satisfied with the bottle packaging. No significant associations were found between HIVTSQc scores and sociodemographic or ART-related variables.

## Introduction

In the last decade, antiretroviral therapy (ART) has demonstrated high efficacy, resulting in higher than 90% viral suppression rates. Therapeutic guidelines identify several regimens that allow adequate long-term suppression of viral load in people living with HIV infection (PLHIV), including single tablet coformulations^1^.

The success of ART depends on treatment satisfaction and adherence, which are impacted by several factors, including medication usage facility, patient perception of results, and side effects. These factors must be considered when making treatment decisions in the clinical management of PLHIV^2^. For all these reasons, satisfaction has been considered relevant in the assessment and differentiation between therapeutic regimens, aiming to improve clinical outcomes, the well-being of PLHIV, and maximizing improvement of health status^3,4^.

Medication packaging in blister packs, with a weekly calendar, has the potential to contribute to improving adherence to ART and optimizing outcomes in the treatment of PLHIV. In the scope of treatment of hypertension, patients with medication in blisters with weekly calendars renewed their prescription more frequently, had higher Medication Possession Ratio (MPR) and better control of blood pressure at 12 months^5^. Additionally, a meta-analysis of 52 studies (n = 22 858) involving patients with different diseases showed an increase in adherence from 63% to 71%, related to the change in the packaging of the medication to blisters with calendars, which demonstrates a practical improvement in treatment adherence because of this tablet package modification^6^.

Biktarvy® is indicated for the treatment of adults infected with human immunodeficiency virus type 1 (HIV-1) without present or past evidence of viral resistance to the integrase inhibitor class, emtricitabine or tenofovir. Currently, Biktarvy® new package contains the same number of units (30 tablets), but packaged in four blisters of seven tablets, with a weekly calendar, and one blister of two tablets. This new packaging has the potential for the patient to improve the adherence to therapy, facilitating the traceability of daily medication dosage, its portability and greater medication discretion, reducing the risk of stigma associated with ART. In Portugal, there was a complete replacement of units of Biktarvy® in bottles by Biktarvy® in blisters, until May 2022^7^.

This project aimed to estimate the impact of the new Biktarvy® packaging containing the same number of units (30 tablets), but packed in four blisters of seven tablets, with a weekly calendar, and one blister of two tablets, on patient’s satisfaction (main outcome) and adherence (secondary outcome) with ART.

## Materials and methods

### Research strategies

The study followed an observational retrospective/prospective design with PLHIV being followed in two Portuguese hospitals. Data collected referred to a period of ART of at least six months before the change of packaging (bottle; retrospective data) and six months of experience with the new packaging (blister; prospective data).

The study was conducted in accordance with the clinical study protocol (CSP), the International Council for Harmonization, Good Clinical Practices, and the Declaration of Helsinki, as well as the applicable European and Portuguese laws and regulations approved by the Ethics Committee of the Centro Académico de Medicina de Lisboa (reference number 226/22).

All patients signed a written informed consent form before entering the study. Each participant received a full explanation of the project goals, procedures, and implied tasks (and burden) from patients. Subjects were informed that their participation was voluntary and assured that they could abandon the study at any time without any prejudice. They also received a copy of the subject’s information and, after being fully clarified, signed the informed consent form.

### Sampling: inclusion and exclusion criteria

The sample size was planned to enroll 100 PLHIV (including at least 20 women) on treatment with Biktarvy®, being over 18 years old, under ambulatory clinical management in two Portuguese hospitals for at least six months and accepting to participate voluntarily in the study.

Patients coming to the hospital pharmacy were included sequentially (non-probabilistic sampling), and the sample size was determined to allow an estimate of the patients with timely prescription delivery in the hospital pharmacy (before the time of prescription collection), with the precision of 7% to 10%. The first patient visit occurred in August 2022, and the last patient visit occurred in December 2023.

### Data collection and instruments

Subjects’ data was directly collected on paper-based clinical reports forms (CRFs). Subsequently, data were transcribed into the study database by a data entry operator. Each patient was given a study reference number, which was archived in the clinical file.

Data collected included demographic information (age, gender, nationality, level of education), clinical and laboratory data (duration of ART, viral load, and CD4 T lymphocytes count). For assessing satisfaction with ART, two instruments have been used: the Portuguese version of the HIV Treatment Satisfaction Questionnaires version “s” (HIVTSQs), at baseline (retrospective assessment related to the previous six months), and the Portuguese version for the HIV Treatment Satisfaction Questionnaires version “c” (HIVTSQc), six months after starting the use of Biktarvy^®^ monthly blisters. HIVTSQs and HIVTSQc have been developed by Woodcock & Bradley, with evidence of sound psychometric properties^8,9^. The HIVTSQs is composed of 10 items being answered in a seven-points Likert-type scale (varying from 0 = “very dissatisfied” to 6 = “very satisfied”). The HIVTSQs provides a global score of satisfaction as well as scores for two subdimensions: the HIVTSQs General Satisfaction/Clinical subscale and the HIVTSQs Lifestyle/Ease subscale; the HIVTSQc also includes a subscale: the HIVTSQc General Satisfaction/Clinical subscale.

Additionally, and for a better understanding of the factors of satisfaction with ART, patients were asked about their level of agreement with four statements, both at baseline and at follow-up: *The packaging of this medication allows me to easily check if I’ve taken my daily pill*, *This medicine comes in a package that lets me take the pill easily and without anyone noticing*, *This medicine’s packaging reminds me that I need to refill my prescription*, and *I don’t like the way this medicine is packaged*. Finally, patients reported their level of agreement with one more statement, at the follow-up moment: *I am more satisfied with this type of medication packaging than with the previous one, which was a plastic bottle*. Level of agreement was indicated through a 5-points Likert scale ranging from *1 = totally disagree* to *5 = totally agree*.

To assess the impact of the new Biktarvy® packaging on adherence, pharmacy medication dispensing was measured, more specifically, by registering the number of tablets supplied in the last delivery per number of days elapsed since the last delivery. Pharmacy medication dispensing data were collected by pharmacy records, and questionnaires were completed while patients waited in the hospital pharmacy (a representation of the study design is presented in Figure 1).

**Figure 1.**
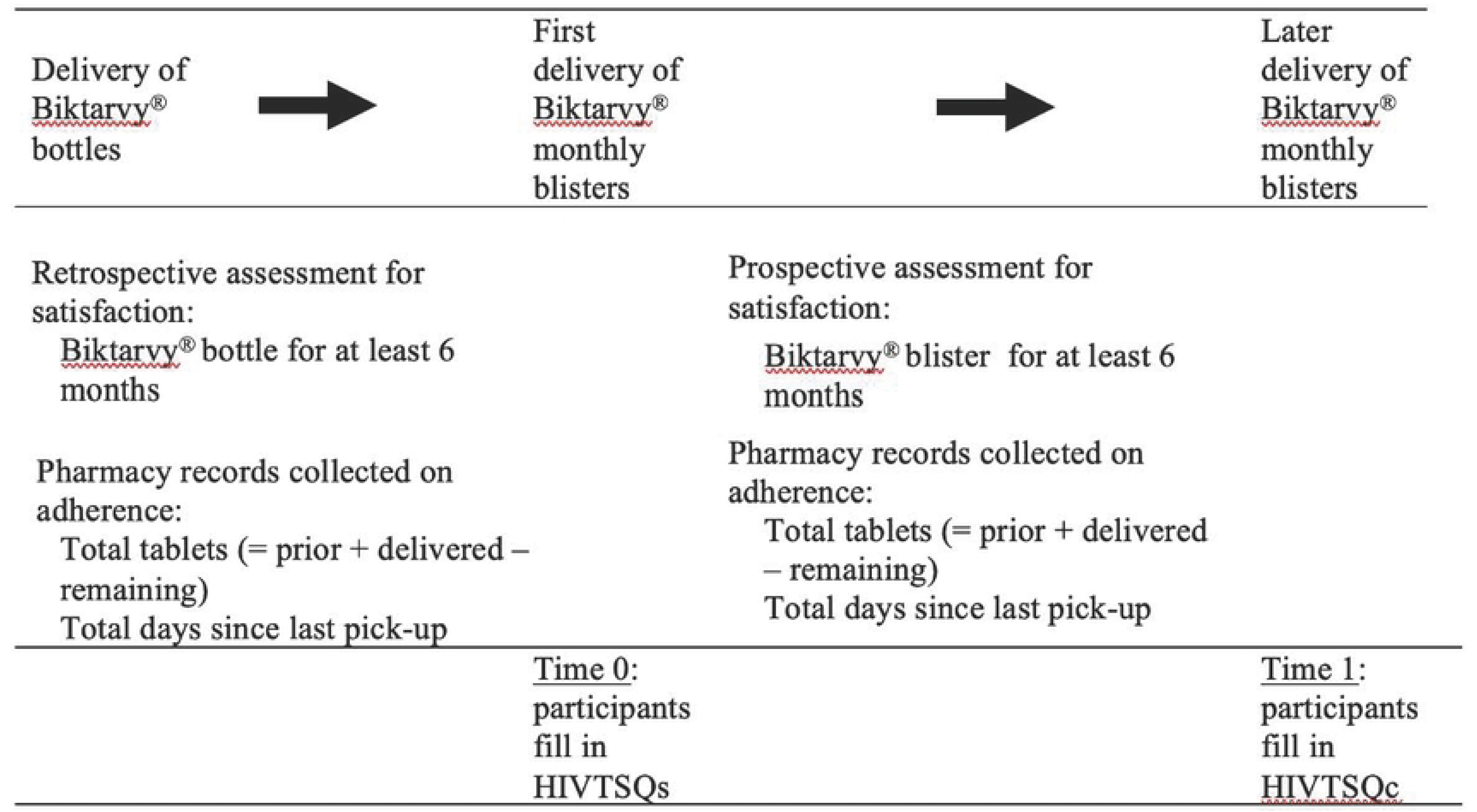
Study design diagram.

### Data analysis

The statistical analyses were conducted using R statistical software version 4.2.1. Descriptive statistics were used to analyze results: percentages and absolute frequencies to describe the categorical variables; minimum, maximum, interquartile range, means, medians, and standard deviation for the quantitative variables.

All ten items from HIVTSQs were summed up to obtain a treatment satisfaction score (0 to 60), where higher scores represent greater satisfaction. The HIVTSQs General Satisfaction / Clinical subscale includes the items 1, 2, 3, 9 and 10 (range: 0 to 30), and the HIVTSQs Lifestyle/Ease subscale includes the remaining items, namely the items 4, 5, 6, 7 and 8 (range: 0 to 30).

For the HIVTSQc, the sum of all items results in a treatment satisfaction (change) score (range: −30 to 30), where positive values are associated with improvement in satisfaction with treatment, the 0 score represents no change, and negative values indicate a deterioration in treatment satisfaction. For the HIVTSQc General Satisfaction / Clinical subscale and HIVTSQc Lifestyle/Ease subscale, the same items as in HIVTSQs subscales were included (range: −15 to 15). Cronbach alpha was used to assess internal consistency for both HIVTSQs and HIVTSQc, and correlations were applied to assess the convergent validity between these two scales.

To evaluate the effect of switching the medication from a standard pill bottle to a monthly blister in patient satisfaction, a unilateral Wilcoxon signed rank test was applied (alternative hypothesis: pseudomedian > 0) to the score in the HIVTSQc scale and its subscales. Differences between independent samples in the distribution of these scores was tested using the Wilcoxon rank sum test.

Linear regression models have been performed to assess the predictive effect of baseline satisfaction on satisfaction change. The scores for HIVTSQc scale, HIVTSQc General Satisfaction subscale, HIVTSQs Clinical subscale, e HIVTSQc Lifestyle/Ease subscale were found as very skewed (left-skewed distribution). Therefore, their values were shifted and reversed for modeling purposes, as follows: *HIVTSQc scale* recoded = 61 + 1 - (*HIVTSQc scale* + 31); *HIVTSQc General Satisfaction / Clinical subscale* recoded = 31 + 1 - (*HIVTSQc General Satisfaction / Clinical subscale* + 16); *HIVTSQc Lifestyle/Ease subscale* recoded = 31 + 1 - (*HIVTSQc Lifestyle/Ease subscale* + 16). Resulting scores were log-transformed and included in the linear regression models as dependent variables. The models included as predictors sex, age group, education, nationality, and antiretroviral therapy duration, adjusting for the treatment satisfaction score at baseline. When studying the predictive effect of treatment satisfaction at baseline (for HIVTSQs scale, HIVTSQs General Satisfaction / Clinical subscale, and HIVTSQs Lifestyle/Ease subscale) on HIVTSQc, HIVTSQc General Satisfaction / Clinical e HIVTSQc Lifestyle/Ease, simple models (no additional variables, in these equations) have been used. The predictors mentioned above were also included all-together in one model (M2). Estimates and 95% confidence intervals (CI), and p-values are presented.

## Results

One hundred and five PLHIV were enrolled in the study. Two were discontinued due to a physician’s decision to switch ART, and one had not completed six months of using Biktarvy^®^ in blisters by the end of the data collection process (therefore, not considered for assessing satisfaction with the blister packaging). Table 1 summarizes the demographic and clinical characteristics of participants at baseline.

**Table 1.**
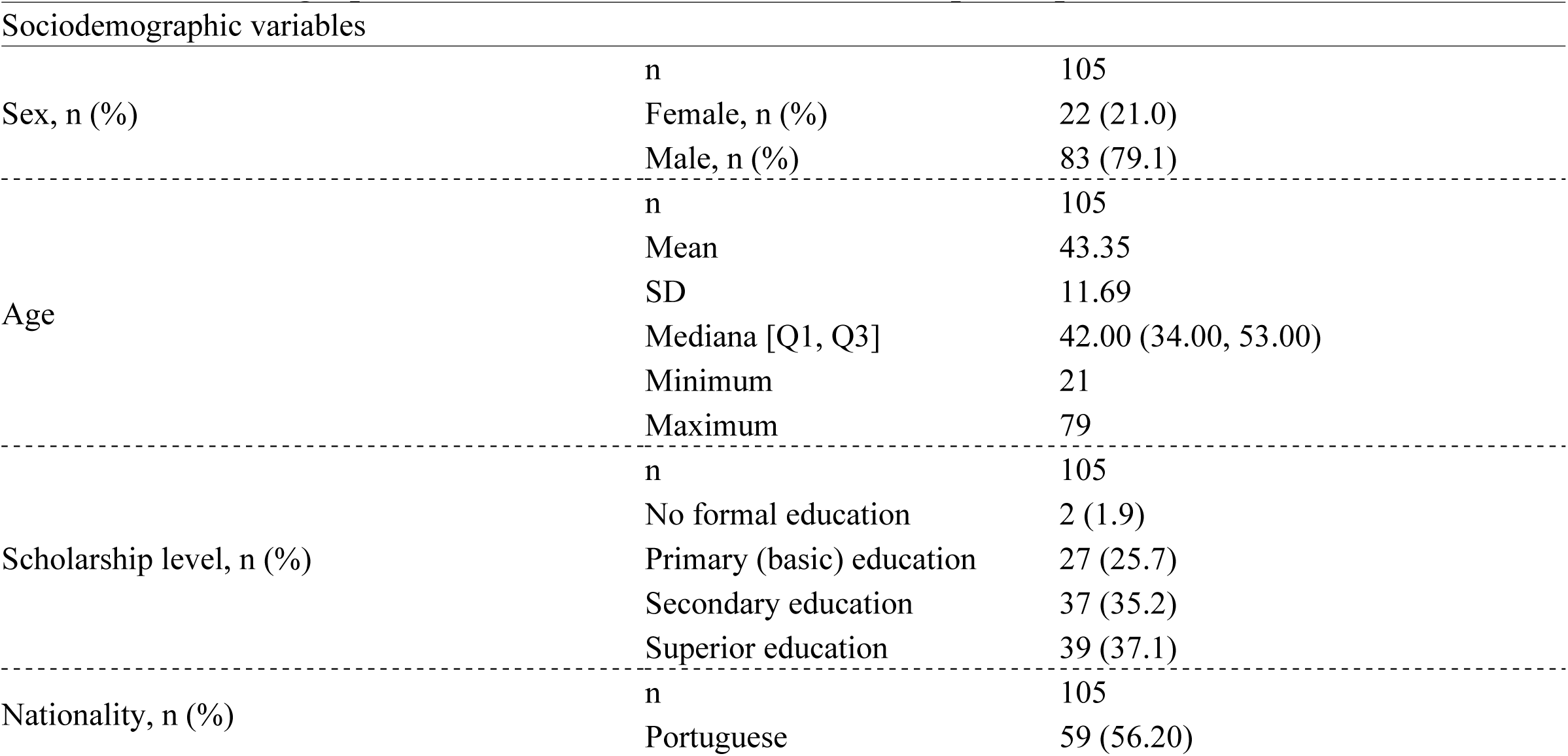
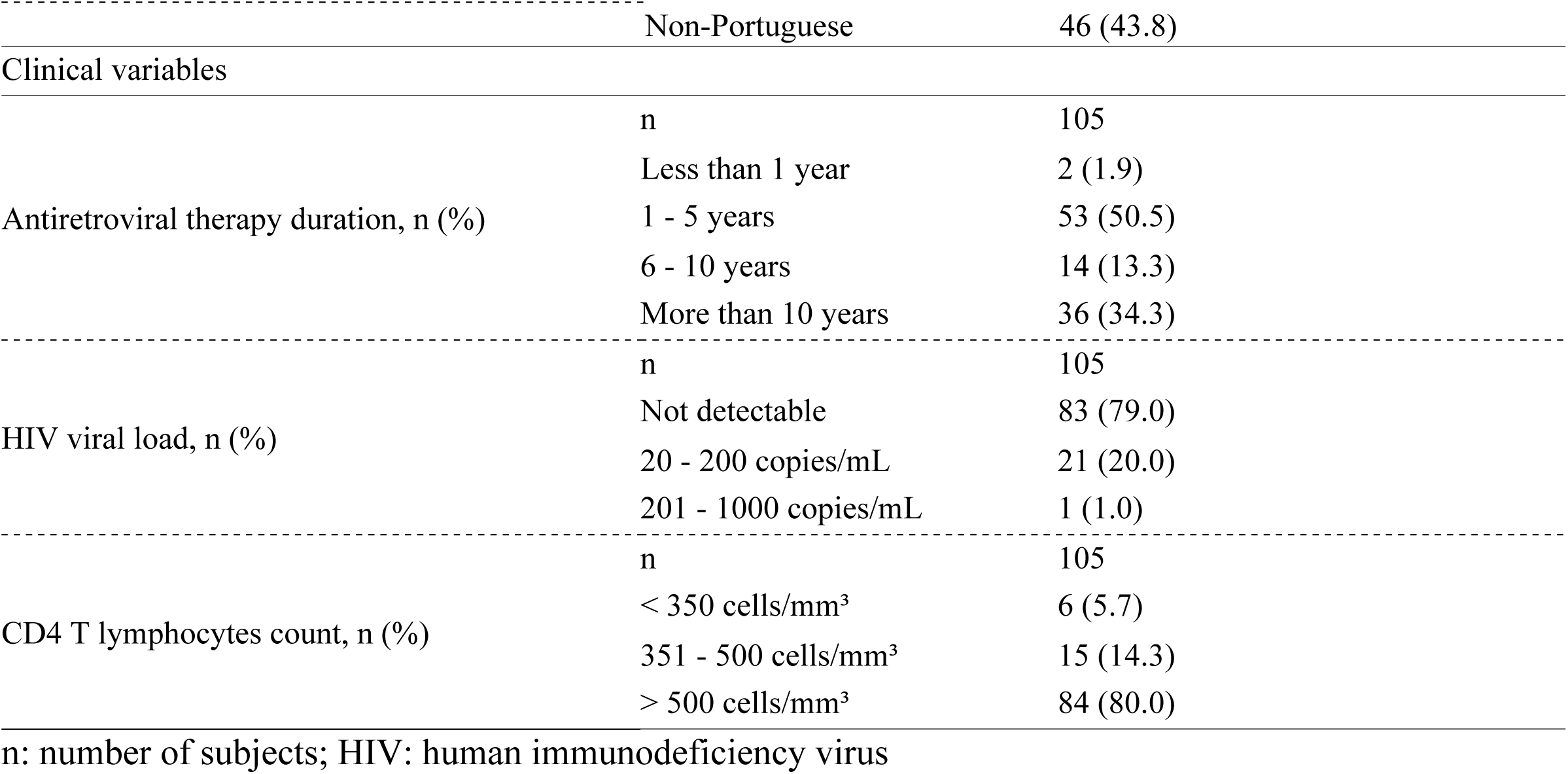
Sociodemographic and clinical characteristics of the participants.

Out of the 105 participants, twenty-one (21.0%) were females. The mean age was 43.35 years (s.d:11.69; min 21, max 79 years old), 35.2% and 37.1% had secondary and superior education, respectively, and 43.8% were born outside Portugal. Only two participants had been following ART for less than one year. 79.0% had a non-detectable viral load, and 80.0% had a CD4T lymphocyte count higher than 500 cells/mm^3^.

The scales used for assessing satisfaction performed well psychometrically: the internal reliability coefficients for HIVTSQs and HIVTSQc were found to be good (alpha = .76 for HIVTSQs; alpha = .88 for HIVTSQc). The correlation between HIVTSQs and HIVTSQc was also significant (rho=.327) (Figure 2).

**Figure 2.**
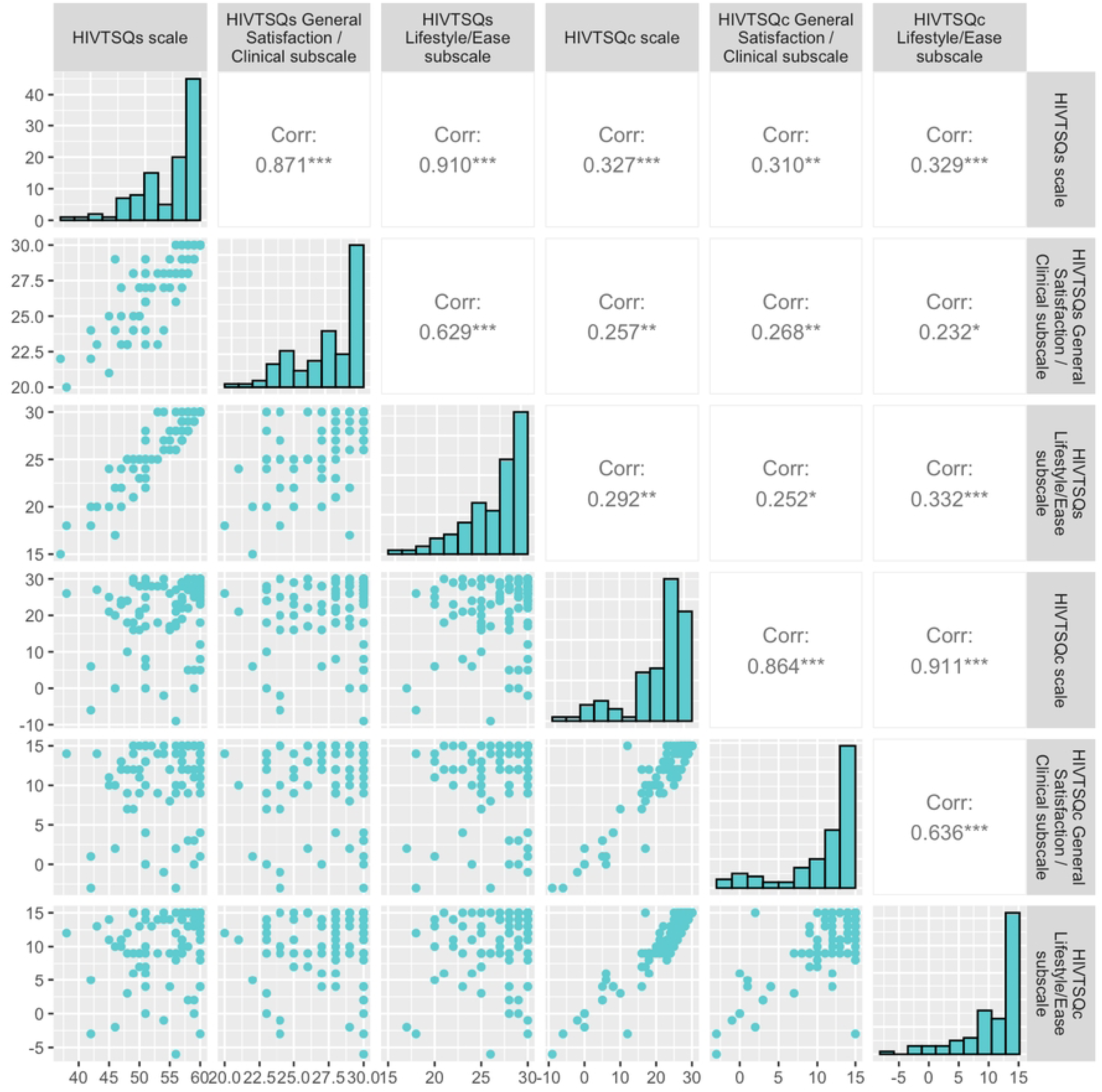
HIVTSQs (and subscales) and HIVTSQc total scores correlations (Spearman correlation)

As Table 2 shows, the scores of HIVTSQs (at baseline) were not found to be associated with sociodemographic variables or the selected clinical indicators. When considering the 6- month assessment (with blister package usage) with HIVTSQc, a significant positive satisfaction evolution was observed (also for the HIVTSQc subscale).

**Table 2.**
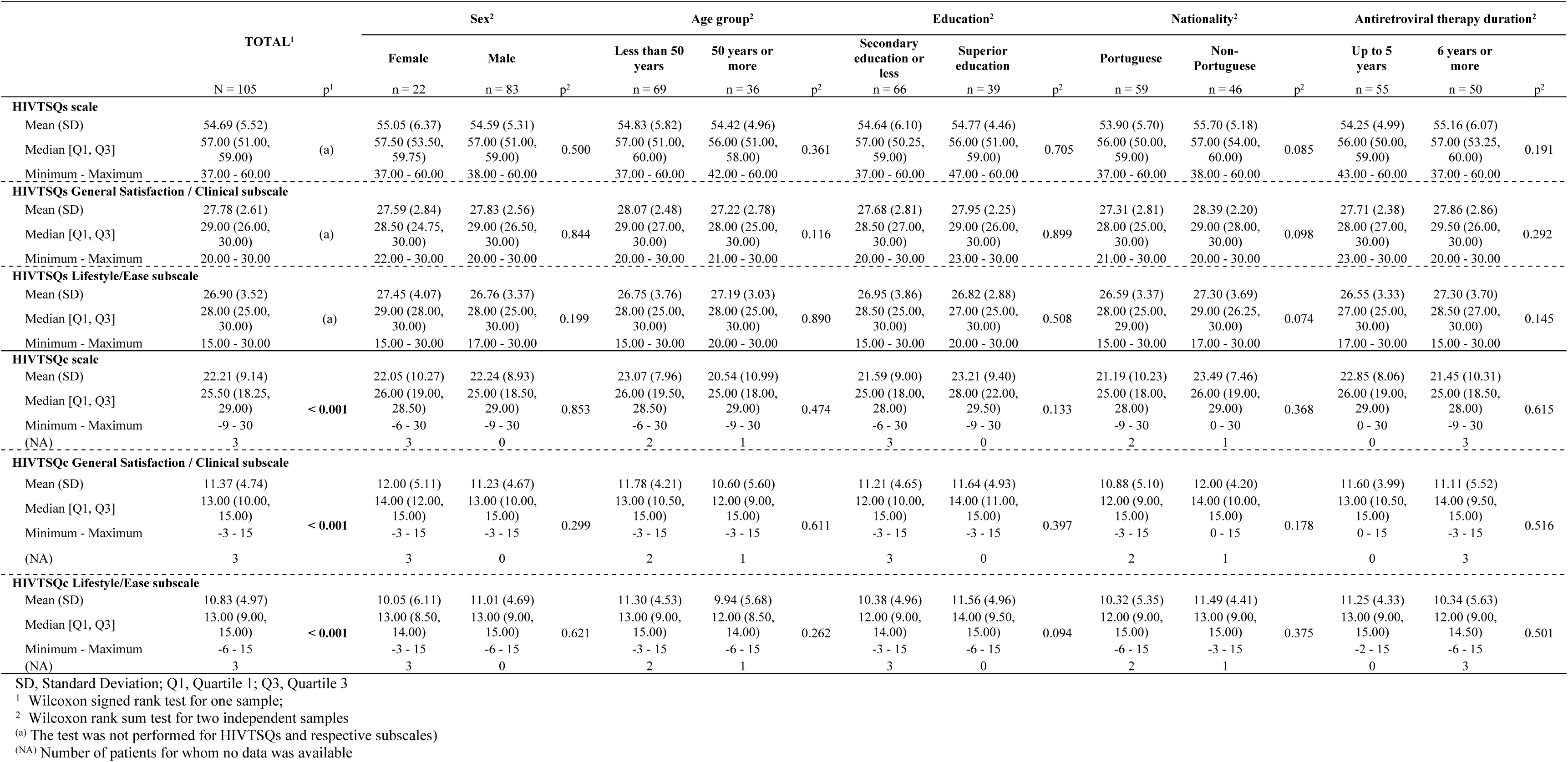
Satisfaction (HIVTSQs and HIVTSQc scores) by sex, age group, education, nationality, and antiretroviral therapy duration.

Table 3 also shows that satisfaction with a blister (measured with HIVTSQc) is not predicted by gender, age group, education level, nationality, or ART duration. On the other hand, an increase in the baseline treatment satisfaction score was associated with a decrease in the treatment satisfaction change score (six months after the switch to blister). For the HIVTSQc scale model, a one-unit increase in the baseline treatment satisfaction score (HIVTSQs) results in a decrease of 6% [exp (0.06)] in the HIVTSQc overall score.

**Table 3.** Satisfaction change (HIVTSQc scores): predictive effect of satisfaction at baseline (HIVTSQs), sociodemografic variables, and clinical variables (linear models)

|  | HIVTSQc scale |  |  |  | HIVTSQc General Satisfaction / Clinical subscale |  |  |  | HIVTSQc Lifestyle/Ease subscale |  |  |  |
| --- | --- | --- | --- | --- | --- | --- | --- | --- | --- | --- | --- | --- |
|  | Model 1 |  | Model 2 |  | Model 1 |  | Model 2 |  | Model 1 |  | Model 2 |  |
|  | b (95% CI) | p | b (95% CI) | p | b (95% CI) | p | b (95% CI) | p | b (95% CI) | p | b (95% CI) | p |
| **HIVTSQs\*** | -0.06 (-0.10; -0.02) | **0.002** | -0.06 (-0.10; -0.02) | **0.003** | -0.10 (-0.17; -0.03) | **0.006** | -0.09 (-0.17; -0.02) | **0.016** | -0.09 (-0.15; -0.04) | **0.001** | -0.10 (-0.15; -0.04) | **<0.001** |
| **Sex** |  |  |  |  |  |  |  |  |  |  |  |  |
| Female | — |  | — |  | — |  | — |  | — |  | — |  |
| Male | -0.03 (-0.56; 0.51) | 0.926 | 0.08 (-0.47; 0.64) | 0.769 | 0.27 (-0.20; 0.74) | 0.252 | 0.29 (-0.20; 0.78) | 0.240 | -0.25 (-0.73; 0.23) | 0.302 | -0.14 (-0.64; 0.35) | 0.559 |
| **Age group** |  |  |  |  |  |  |  |  |  |  |  |  |
| Less than 50 years | — |  | — |  | — |  | — |  | — |  | — |  |
| 50 years or more | 0.12 (-0.32; 0.56) | 0.591 | 0.03 (-0.44; 0.49) | 0.913 | 0.02 (-0.37; 0.41) | 0.918 | 0.02 (-0.39; 0.44) | 0.914 | 0.26 (-0.12; 0.65) | 0.178 | 0.17 (-0.23; 0.57) | 0.409 |
| **Education** |  |  |  |  |  |  |  |  |  |  |  |  |
| Secondary education or less | — |  | — |  | — |  | — |  | — |  | — |  |
| Superior education | -0.34 (-0.76; 0.08) | 0.114 | -0.31 (-0.76; 0.15) | 0.183 | -0.13 (-0.51; 0.25) | 0.498 | -0.17 (-0.57; 0.23) | 0.390 | -0.36 (-0.73; 0.01) | 0.060 | -0.27 (-0.67; 0.12) | 0.174 |
| **Nationality** |  |  |  |  |  |  |  |  |  |  |  |  |
| Portuguese | — |  | — |  | — |  | — |  | — |  | — |  |
| Non-Portuguese | -0.10 (-0.52; 0.33) | 0.655 | -0.07 (-0.50; 0.35) | 0.735 | -0.16 (-0.54; 0.21) | 0.388 | -0.16 (-0.54; 0.23) | 0.420 | -0.15 (-0.52; 0.23) | 0.435 | -0.11 (-0.49; 0.26) | 0.549 |
| **Antiretroviral therapy duration** |  |  |  |  |  |  |  |  |  |  |  |  |
| Up to 5 years | — |  | — |  | — |  | — |  | — |  | — |  |
| 6 years or more | 0.21 (-0.21; 0.62) | 0.331 | 0.14 (-0.31; 0.59) | 0.541 | -0.08 (-0.45; 0.29) | 0.670 | -0.08 (-0.48; 0.32) | 0.705 | 0.24 (-0.13; 0.61) | 0.207 | 0.11 (-0.28; 0.51) | 0.574 |

CI, Confidence Interval
\* These coefficients report to scores in HIVTSQs scale, HIVTSQs General Satisfaction / Clinical subscale and HIVTSQs Lifestyle/Ease subscale in models for HIVTSQc scale, HIVTSQc General Satisfaction / Clinical subscale and HIVTSQc Lifestyle/Ease subscale, respectively. For analysis, HIVTSQc scores were shifted, reversed and log transformed.
Model 1, Individual models adjusted for each variable presented, plus the HIVTSQs score
Model 2, Model adjusted for all the variables presented

This predictive value of baseline satisfaction (HIVTSQs) was also observed for the HIVTSQc subscale (general satisfaction/clinical subscale).

Plots in Figure 2 provide some insight into the facets of blister packaging, as perceived by patients (compared with bottles) that may support the observed positive evolution of satisfaction. Although no significant difference was found in the proportion of patients reporting appreciating positively the packaging (bottle and blister), three main facets of usage were found to be better considered by patients: a significantly higher proportion of them considered that the blister helps them to keep track of daily intake of pills; a higher proportion indicate that blister packaging usage is more discrete than bottle usage; and a higher proportion report that blister packaging helps them to recall the refill momentum.

Overall, and as seen in Figure 3, 68.6% of patients report being more satisfied with the blister packaging than with bottle packaging.

**Figure 3.**
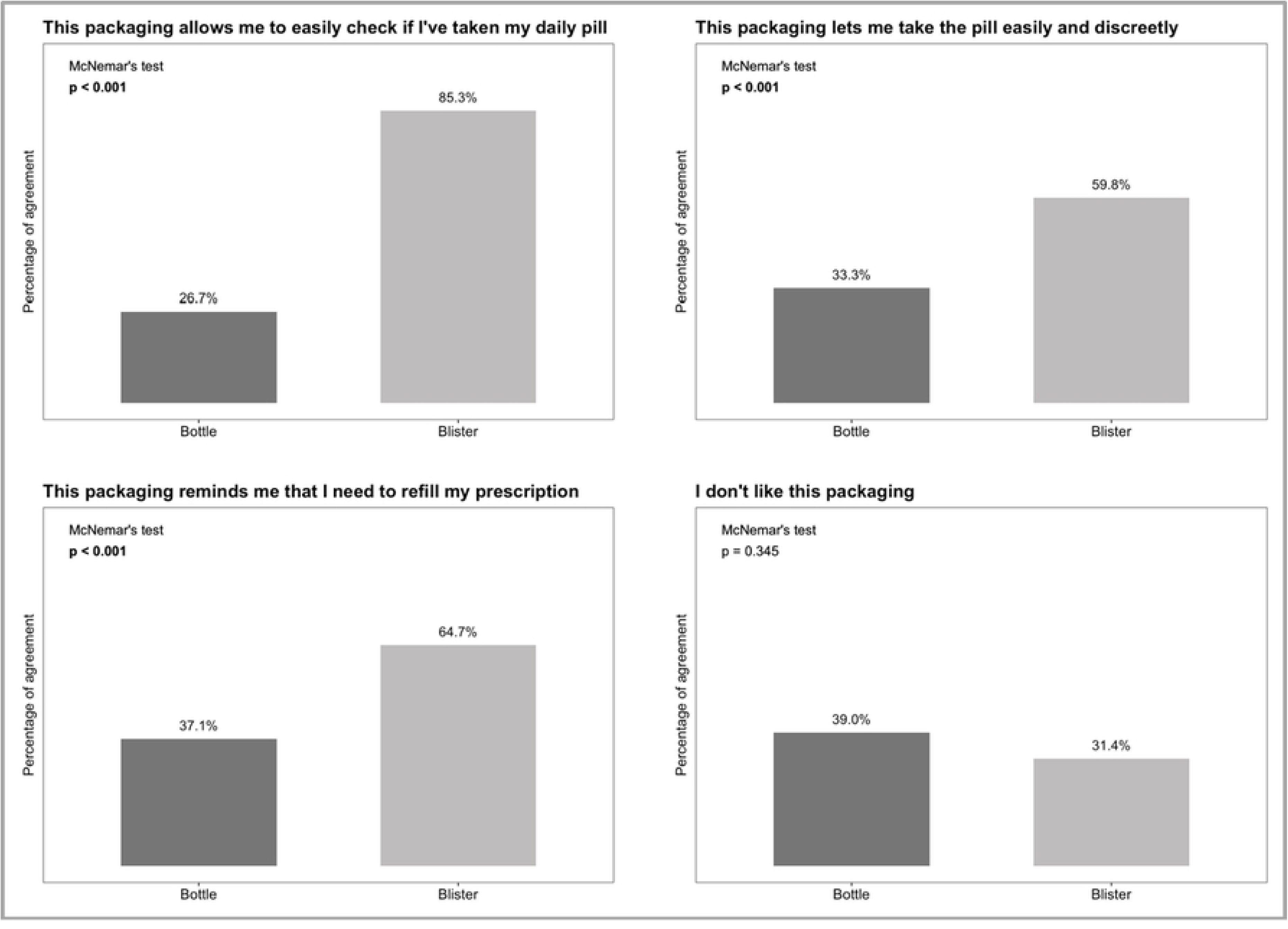
Frequencies of item response on the specifically developed questionnaire between packaging methods.

**Figure 4.**
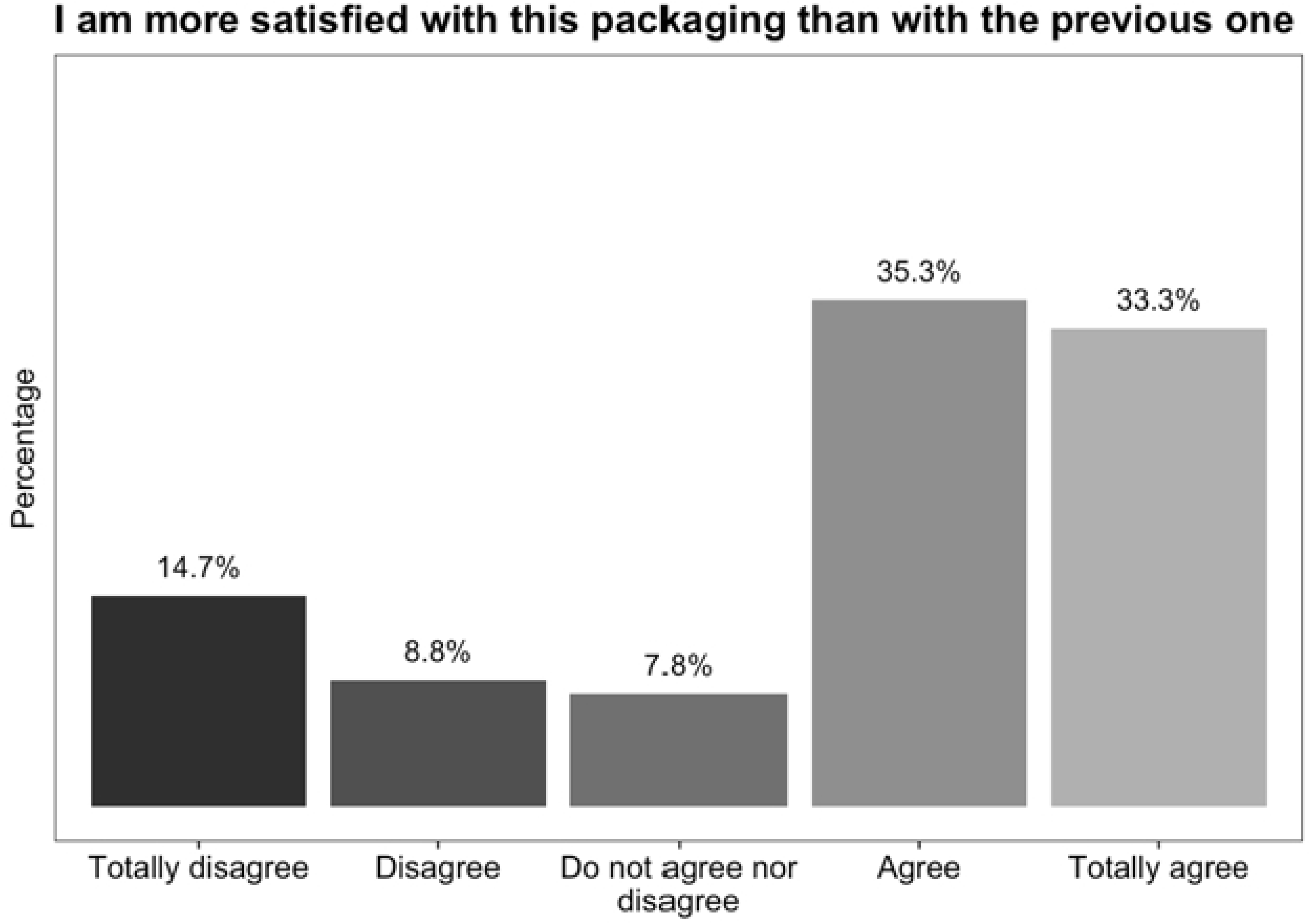
Response distribution on patients’ satisfaction with blister packaging, when compared with bottle packaging.

Regarding adherence, no significant differences were observed between the standard pill bottle and monthly blister packaging: 89.7% and 91.3% of adherence, respectively.

## Discussion

Main aim of this project was to assess if introducing a new packaging would affect satisfaction and adherence with ART. The study was planned as a two-center, observational retrospective and prospective study with 18+ years old PLHIV conducted in Portuguese hospitals (non-random sampling). The bottle effect was assessed retrospectively for at least six months, and the blister effect was prospectively evaluated for a period of at least six months of usage. Overall, 105 PLHIV were enrolled and 102 completed the prospective component of the study.

The main outcome of the study was satisfaction with ART treatment. Patients reported an increase in satisfaction with the Biktarvy^®^ blister packaging (compared with previous bottle packaging). And this increase in satisfaction was independent of several main sociodemographic and clinical characteristics. Importantly, the patients who became more satisfied with ART treatment (when using blister packaging) were the least satisfied with bottle packaging.

Another main finding regards the perception of utility from the blister packaging: after a minimum of six month of experience, the blister (which includes a weekly calendar) was considered by the majority (85%) of patients as an effective tool to keep track of their daily intake of pills (significant higher percentage than the appreciation for bottle packaging), and about two third of patients considered that it is helpful as a reminder of the adequate moment of refilling.

## Conclusion

To our knowledge this is the first study evaluating satisfaction with blister calendar package usage of ART by PLHIV, and the same for medication packaging in blister packs with a weekly calendar for the treatment of different diseases.

Overall, these findings support the advantages of the new packaging. Nevertheless, adherence was not found to be significantly higher for blister than for bottle packaging (though with a slight increase of proportion in adherence: 1.6 percentual points). This issue calls for future research, also because the lapse of time after introducing the new packaging was limited (about six months). Larger samples, including selected populations with more extended periods of blister utilization, may provide sounder evidence about the adherence potential of this Biktarvy^®^ new packaging.

## Data Availability

All relevant data are within the manuscript and its Supporting Information files.

## Acknowledgments

The authors thank the patients who accepted to participate in the study, Prof. Clare Bradley and Health Psychology Research Limited (HPR) for the HIVTSQs and HIVTSQc, and Ana Sequeira and Hugo Caldeira for technical assistance.

